# Immunogenicity, Safety and Effectiveness of COVID-19 Pfizer-BioNTech (BNT162b2) mRNA Vaccination in Immunocompromised Adolescents and Young Adults: A systematic Review and Meta-Analyses

**DOI:** 10.1101/2023.01.20.23284812

**Authors:** Patrick DMC Katoto, Mireille AM Kakubu, Jacques L. Tamuzi, Amanda S. Brand, Adaeze Ayuk, Liliane N. Byamungu, Charles S. Wiysonge, Glenda Gray

## Abstract

People with weak immune systems are more likely to develop severe COVID-19, less likely to be included in vaccine controlled studies but more likely to be under-vaccinated. We review post-marketing studies to examine the immunogenicity, safety and effectiveness of BNT162b2 vaccine in immunocompromised adolescents and young adults (AYA). We searched more than three international databases from 2020 to 30 May 2022 and used the ROBINS-I for bias assessment. Random effect model was used to estimate pooled proportion, log RR, and mean difference. Egger’s regression and Begg’s rank correlation were used to examine publication bias. 47 full texts were reviewed, and nine were included. Conditions studied were rheumatic diseases, diabetes mellitus, Down syndrome, solid tumours, neurodisability, and cystic fibrosis. Eight studies used cohort designs and one used cross-sectional designs. Europe led most of the investigations. Most studies had unclear risk of bias and none could rule out selection bias, ascertainment bias, or selective outcome reporting. The overall estimated proportion of combined local and systemic reactions after the first BNT162b2 vaccination was 30%[95% CI: 17-42%] and slightly rose to 32% [95% CI: 19-44%] after the second dose. Rheumatic illnesses had the highest rate of AEFI (40%[95% CI: 16-65%]), while cystic fibrosis had the lowest (27%[95% CI: 17%-38%]). Hospitalizations for AEFIs were rare. Healthy controls exhibited higher levels of neutralizing antibodies and measured IgG than immunocompromised AYA, although pooled estimations did not demonstrate a statistically significant difference after primary dose. BNT162b2 is safe and effective in immunocompromised AYA, with no significant difference to healthy controls. However, current evidence is low to moderate due to high RoB. Our research advocates for improving methodology in studies including specific AYA population.

## Introduction

The severe acute respiratory syndrome (SARS) pandemic caused by the SARS-coronavirus-19 type 2 (SARS-CoV-2) has resulted in about (as of 5:01pm CET, 11 November 2022) 630,832,131 confirmed cases of COVID-19, including 6,584,104 deaths, reported to WHO[1]. The pandemic has afflicted all population groups, including children, adolescents, and young adults (AYA), despite their milder course[2]. Children and AYA with comorbidities such as chronic lung disorders, obesity, cardiovascular disease, kidney disease, solid organ transplant, malignancies, or rheumatic diseases, as demonstrated in adult populations, have a higher risk of severe diseases with outcomes such as multi-system inflammatory syndrome in children (MIS-C), intensive care unit (ICU) or life support needs, and increased hazards of death[3–7]. Furthermore, COVID-19 clinical presentation in AYA with comorbidities is uncommon and may not follow the conventional epidemiological profile. AYA with comorbidities, for example, may acquire MIS-C even after a moderate illness, especially if they have already been exposed to an infected adult[8,9]. AYA with Down syndrome face an additional risk due to their low immunity, putting them at risk of getting serious COVID-19-related disorders[10]. Similarly, certain comorbidities may expose AYA to a more severe form of SARS-CoV-2 than others. According to global cancer data, 20% of children with cancer acquire severe SARS-CoV-2 infection[9].

Vaccination is a well-established method for preventing severe disease development throughout the life course, particularly in persons with chronic and immune-compromising medical illnesses or at increased risk of infection owing to immunosenescence. The flu vaccine, for example, is associated with a significant reduction in the risk of hospitalization for influenza illnesses in a predominantly elderly population[11], and the Haemophilus influenzae type b (Hib) vaccine has significantly improved the survival of children with sickle cell disease living in low-income countries[12]. To combat the COVID-19 pandemic and its harmful consequences, safe and effective vaccines were developed and licensed first for adults, and subsequently approved for AYA under the age of 16. In AYA, messenger-RNA (m-RNA) vaccines such as BNT162b2 (Pfizer-BioNTech) that encode the SARS-CoV-2 full-length spike responsible for severe acute respiratory syndrome are commonly employed. As of 8 November 2022, a total of 12,885,748,541 vaccine doses have been administered[1]. Serious adverse effects were uncommon in randomised clinical trials (RCTs) involving AYA participants, and vaccination effectiveness was near to 100 percent[13,14]. RCT data, on the other hand, are less likely to match real-world data as people with comorbidities are much less likely to be enrolled in RCTs. Furthermore, in post-marketing studies, the BNT162b2 vaccine was associated to adverse events following immunization (AEFI) not recorded in RCTs, such as myocarditis[15], and vaccination effectiveness differed from that observed in RCTs[16]. It is also recognized that vaccine-preventable diseases are more likely to be severe in people with immunocompromised conditions, and that these people are more likely to be hesitant to receive vaccines. Unfortunately, low immunization rates in these categories are also caused by healthcare practitioners’ fails to implement recommendations[17].

Considering the paucity of data in immunocompromised AYA, we thoroughly examined the evidence from post-marketing surveillance to assess the effectiveness, safety, and tolerability of the BNT162b2 vaccine against SARS-CoV-2 in AYA to enable for data-driven policy decision making.

## Materials and Methods

### Criteria for considering studies for this review

#### Types of studies

Non-randomized interventional studies (post-authorization surveillance data), independent of method or unit of allocation, were included, as were observational studies, such as cohort studies (both prospective and retrospective), case-control studies, and cross-sectional studies. Participants in the included studies could be followed up on for any length of time. We considered studies that included a subset of eligible participants (e.g., children and adolescents) if the results for the eligible subset of participants were published separately. If this was not possible, such studies were included if 90% or more of the sample was AYA. We omitted reviews, case series, and case reports, as well as non-human subject studies. Studies involving children under the age of ten or individuals above the age of twenty-four were also barred.

#### Types of interventions

We included studies investigating any injectable BNT162b2 vaccine intended for the prevention, or mitigation of symptoms, of SARS-CoV-2-infection. All studies involving one or more primary doses (usually doses 1 or 2) and boosters of the BNT162b2 vaccine were eligible for inclusion. Studies were included if interventions were compared with or without a placebo vaccine or with another SARS-CoV-2 vaccine. Studies reporting on different COVID-19 vaccinations only or predominantly were also excluded (mRNA1273, CoronaVac, etc.).

#### Types of outcomes

The outcomes of interest were broadly classified as vaccination i) safety profile (defined in supplemental table 1), ii) tolerability, and iii) efficacy or effectiveness.

### Search methods for the identification of studies

We used a comprehensive search strategy designed to identify the maximum number of eligible studies regardless of language or publication status within a restrictive timeframe. To maximize sensitivity, the techniques did not differentiate between “safety” and “efficacy or effectiveness”. Records were identified through a systematic search of MEDLINE (PubMed), Embase (Ovid), Web of Science and Cochrane library (CENTRAL). The search was initially designed for MEDLINE (PubMed), but it has since been adapted for use with other sources. PubMed search approach is detailed in **box 1** of the supplemental materials. Other sources such as references of included studies, the WHO database, and the Centres for Disease Control and Prevention (CDC) website as well as the Advisory Committee on Immunization Practices (ACIP) meetings were also searched to identify any references to additional studies that could be potentially eligible. The search began on February 15, 2022, and proceeded on the 15th of each month until May 30, 2022.

### Data collection and analysis

#### Selection of studies

Covidence software (Veritas Health Innovation, Melbourne, Australia) was used to import all search results. For de-duplication and screening methods, see www.covidence.org. To select potentially relevant full studies, two reviewers independently examined titles and abstracts. Any disagreements between reviewers were handled through dialogue, with a third reviewer making the final decision. For each record deemed possibly eligible, full text reports were obtained. Two reviewers separately screened these full texts to identify research for inclusion in the review, with any disagreements resolved by consensus discussion with the assistance of a third reviewer. The rationale for not including full-text reports was documented. When a full report of a study (e.g., conference proceedings) was not accessible and there was insufficient material for inclusion, the study was labeled as ‘ awaiting categorization’ until the next search update. Preprint papers were not included unless they were peer reviewed at the time of the last search. A PRISMA flow diagram was used to document and summarize the flow of experiments (**Figure 1**).

**Figure 1:**
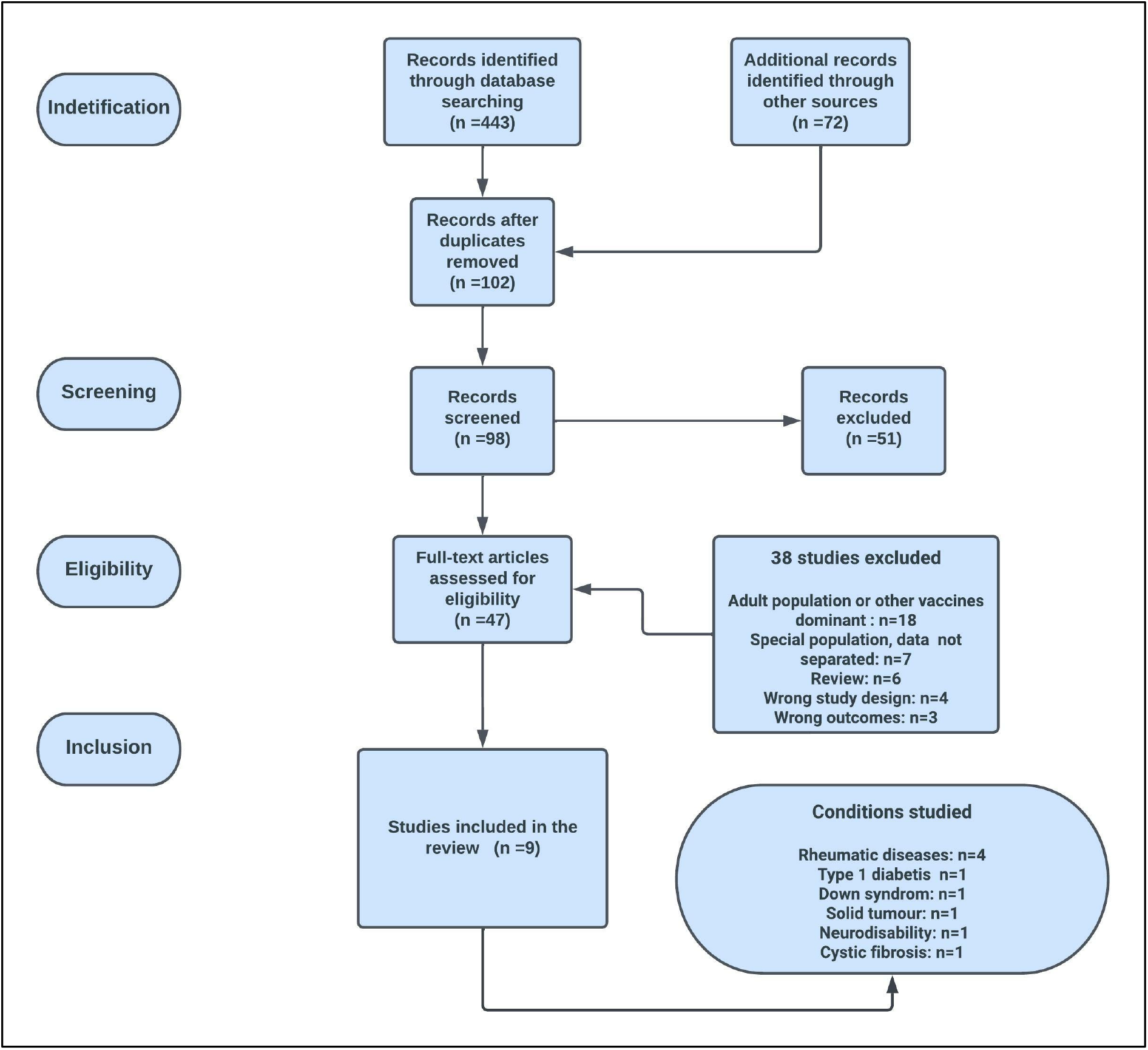
Study PRISMA flow chart.

#### Data extraction and management

A single reviewer extracted study characteristics and outcomes data, which was then crosschecked by a second reviewer to ensure consistency. The extractions were conducted in Covidence utilizing a pre-piloted extraction form. A table of characteristics of included studies was created utilizing descriptive information extracted from studies and exported from Covidence into an excel document.

#### Assessment of risk of bias in included studies

Two reviewers assessed the risk of bias (RoB) of included studies using the Cochrane RoB tool[18] for randomized evidence, and the ROBINS-I tool[19] for non-randomized studies. One senior reviewer conducted all RoB assessments as the independent duplicate reviewer. Standard domains for each tool were used, and an overall risk of bias was determined using a worst-domain scenario approach. As a result, we judged the overall risk of bias for each study as ‘ low risk’, ‘ unclear risk’ or ‘ high risk’ and reported the main reason in the summary table for RoB. Any discrepancies in individual domain judgments as well as overall judgements were resolved through consensus or adjudication by a third review author.

### Statistical analysis

Stata 16.0 was used to conduct the meta-analyses (Stata corp., College Station, TX, USA). To account for study heterogeneity, the random effects model estimated pooled proportion, log RR, and mean difference with 95 % confidence intervals (CI). To quantify heterogeneity, I^2^ statistics were utilized. All analyses were initially performed in comparison to the administration of the first and second doses of BNT162b2 vaccines. Subgroup analyses were then performed, which comprised rheumatic illnesses, severe neurodisabilities, Down syndrome, type 1 diabetes, cystic fibrosis, and solid tumours. Finally, the combined effect of the first and second doe was investigated. When the standard error (SE) in a study was not published, we determined it from the proportion using the formula: SE = P (1-P)/N and 95 % CI = P 1.96 SE, where P was the proportion and N was the sample size comprising immunocompromised adolescents[20]. Studies that reported the median and IQR were converted to mean (SE)[21], and BAU/ml or U/ml to AU/ml using online computations. A meta-regression was also performed on the local and systemic reactions. Because the review comprised less than ten studies from all forest plots, Egger’ s regression analysis and Begg’ s rank correlation analysis were used to investigate the probability of publication bias.

## Results

### Characteristic of included studies

Of the 443 records found in the various data bases analyzed, 47 full texts were carefully reviewed for eligibility, and nine studies were included (**Figure 1**). Rheumatic disorders were studied in four studies, and type 1 diabetes, Down syndrome, solid tumor, neurodisability, and cystic fibrosis were studied in one each. Eight studies (Akgün 2022; Dimopoulou 2022; Heshin-Bekenstein 2022; King 2022; Michos 2022; Piccini 2022; Riviere 2021; Valentini 2022)[3,4,8–10,22–24] employed a cohort design, whereas one used a cross-sectional design(Haslak 2022)[6]. Most investigations were undertaken in Europe (**Supplementary Table 2**).

### Risk of bias

The assessment of RoB for these studies was conducted using ROBINS-I (**Supplementary Table 3**). Six studies had an unclear overall risk of bias (Akgün 2022; Dimopoulou 2022; Haslak 2022; King 2022; Piccini 2022; Riviere 2021). None of these studies could rule out selection bias, ascertainment bias, or selective outcome reporting. Bias due to confounding could also not be ruled out in four studies (Akgün 2022; King 2022; Piccini 2022; Riviere 2021); while bias in the classification of interventions could not be ruled out in two (Haslak 2022; Piccini 2022); deviations from intended interventions could not be ruled out in five (Akgün 2022; Dimopoulou 2022; King 2022; Piccini 2022; Riviere 2021); and attrition bias could not be ruled out in four (Akgün 2022; Dimopoulou 2022; King 2022; Piccini 2022). Furthermore, three (Akgün 2022; Haslak 2022; Riviere 2021) had other sources of bias identified – this related mostly to small sample sizes and short post-vaccination follow-up.

Three studies had a high overall risk of bias (Heshin-Bekenstein 2022; Michos 2022; Valentini 2022). Three studies were at high risk of bias due to confounding (Heshin-Bekenstein 2022; Michos 2022; Valentini 2022); one was at high risk of attrition bias (Heshin-Bekenstein 2022); and one was at high risk of ascertainment bias (Heshin-Bekenstein 2022). Selection bias, bias due to deviations from intended interventions and selective outcome reporting could not be ruled out in any of these studies; bias in the classification of interventions could not be ruled out in two studies (Michos 2022; Valentini 2022), attrition bias could not be ruled out in one (Michos 2022), and ascertainment bias could not be ruled out in two studies (Michos 2022; Valentini 2022).

### Any local AEFI

**Figure 2A** shows the tolerability of dose 1 BNT162b2 vaccine among adolescents. Immunocompromised conditions include rheumatic diseases, severe neurodisabilities, Down syndrome, type 1 diabetes, and solid tumours, with an overall pooled proportion of any local reaction of 28%[95% CI: 11%-44%]. The following local reactions were observed among the six studies included in the pooled results: pain, swelling, itching, and erythema. The subgroup analysis revealed that diabetes mellitus type 1 adolescents had the highest rate of any local proportion reactions 72%[95%CI: 56%-87%], solid tumours (46%[95%CI: 19%-73%], rheumatic diseases (27%[95%CI: 4%-50%], down syndrome (13%[95%CI: −9%-35%], and severe neurodisabilities 4%[95%CI: −4%-11%]. The summary effect or pooled proportional estimate of any local reaction following the second dose of BNT162b2 vaccine among immunocompromised adolescents was found to be 23% [95% CI: 10%-37%]. Pain, swelling, erythema, itching, and pruritus were the most common local reactions following dose 2 of the BNT162b2 vaccine. Adolescents with type 1 diabetes had the highest pooled estimate of any local reaction (64% [95%CI: 49%-79%], followed by rheumatic diseases (23% [95%CI: 6%-41%], solid tumours 15% [95%CI: −4%-35%], Down syndrome 14% [95%CI: −5%-34%], and severe neurodisabilities 9% [95%CI: −0.3%-21%] (**Figure 2B**).

**Figure 2:**
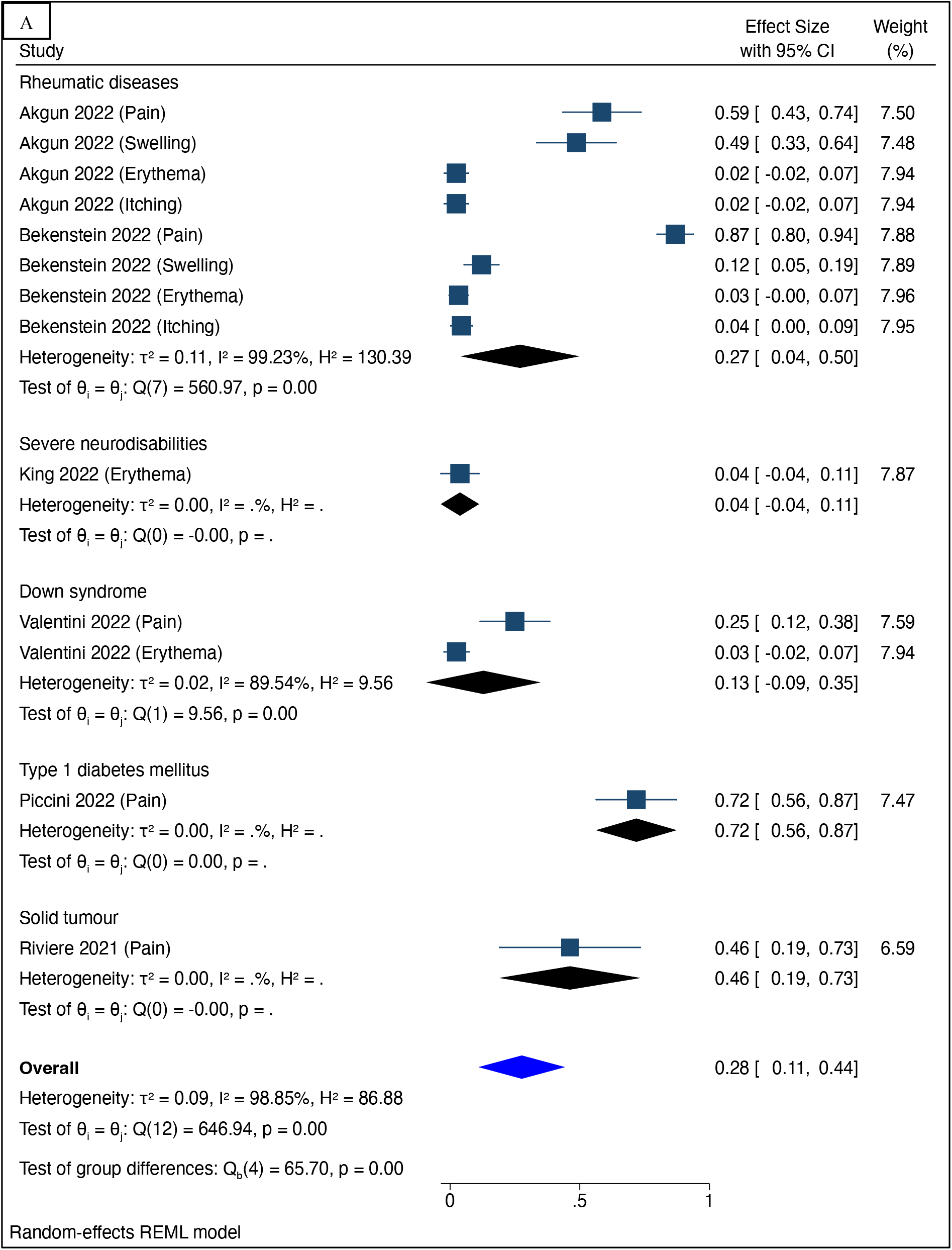
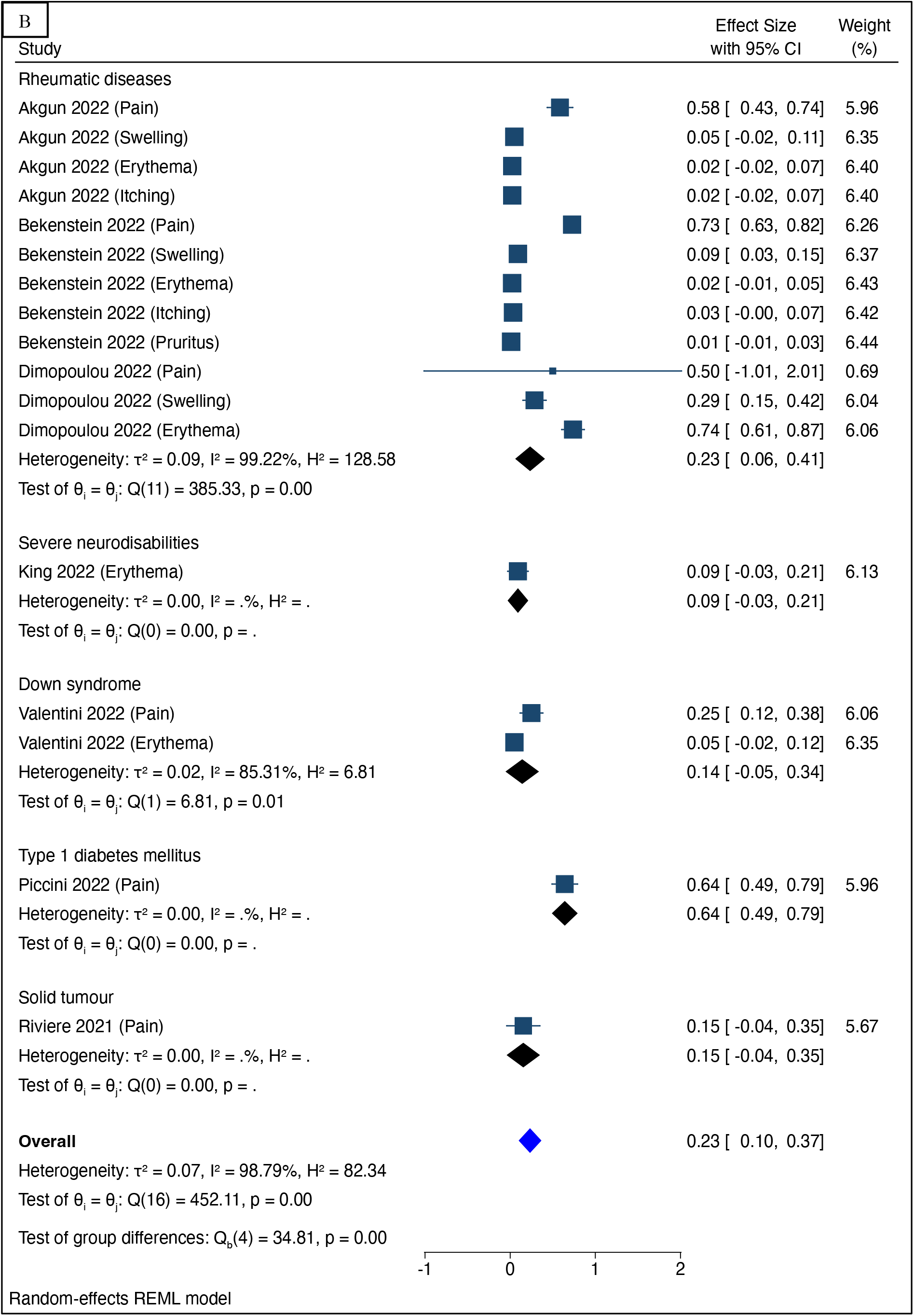
Forest plot of the proportion of any local reaction among immune-compromised adolescents following receipt of the first (Panel 2A) and second (Panel 2B) doses of BNT162b2 vaccine.

### Any systemic AEFI

The most common systemic reactions after dose 1 in the meta-analysis evaluating any systemic reaction following administration of BNT162b2 vaccine in immunocompromised adolescents were fever, muscle ache, headache, fatigue, running nose, joints pain, chills, feeling unwell, hospitalization, weakness, exacerbation, nausea, and vomiting, with an overall pooled proportion of 9%[95%CI: 6%-11]. Adolescents with diabetes mellitus and those with severe neurodisabilities had the highest incidence of systemic reactions, at 20%[95% CI: 6%-32%] and 20%[95% CI: 6%-36%], respectively. The proportion of adolescents with systemic reactions among those with solid tumour was 12% [95%CI: 2%-22%], while those with rheumatic diseases and Down syndrome had 6% [95%CI: 5%-10%] and 3% [95%CI: 1%-6%], respectively (**Figure 3A**).

**Figure 3:**
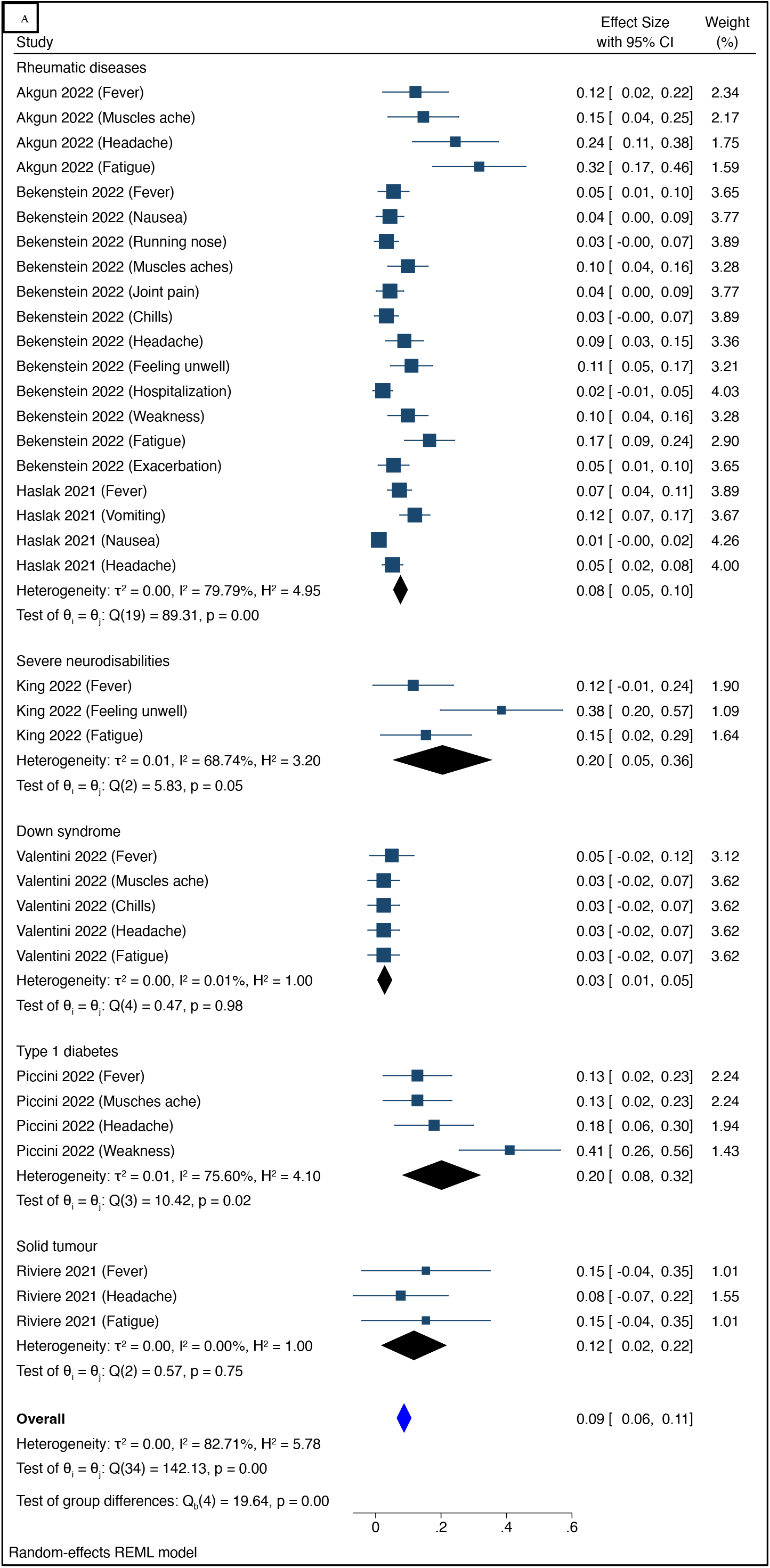
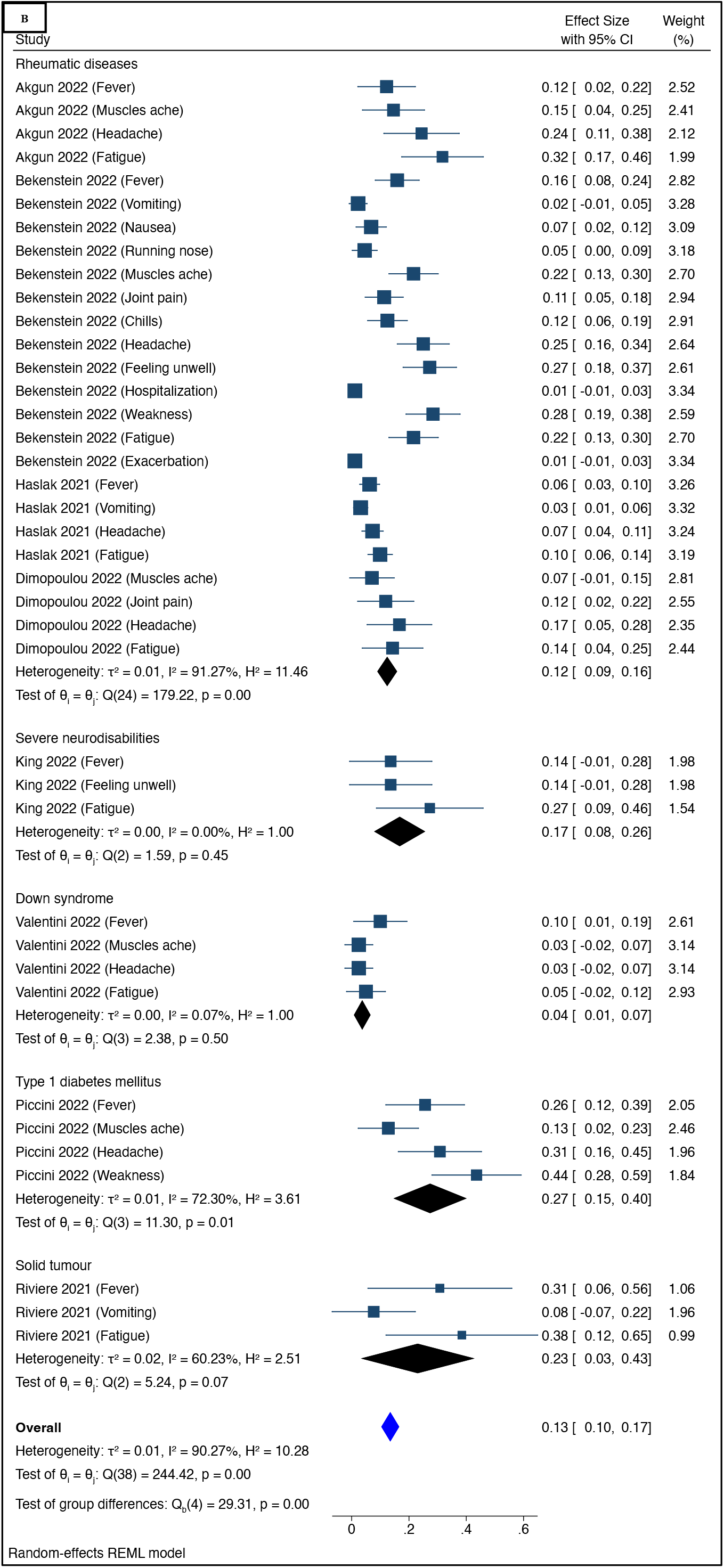
Forest plot of the proportion of any systemic reaction among immune-compromised adolescents following receipt of the first (Panel 2A) and second (Panel 2B) doses of BNT162b2 vaccine.

The overall pooled proportion of any systemic reaction after dose 2 of BNT162b2 vaccine was 13% [95% CI: 10%-17%]. Fever, muscle ache, headache, fatigue, running nose, joint pain, chills, feeling unwell, hospitalization, weakness, exacerbation, nausea, and vomiting were among the common systemic reactions observed. As with the first dose, adolescents with diabetes mellitus exhibited the highest proportion of systemic reactions 27%[95%CI: 15%-40%], followed by those with solid tumour 23%[95%CI: 3%-43%], severe neurodisabilities 17%[95%CI: 6%-20%] then by those with rheumatic diseases 12%[95%CI: 9%-16%] and Down syndrome 4% [95% CI: 1%-7%] (**Figure 3B**).

### Any local and systemic AEFI

The overall proportion of combined AEFIs in immunocompromised adolescents in studies that reported on combined systemic and local AEFIs following the first dose of BNT162b2 vaccination was 30%[% CI: 17% − 42%]. A high proportion of AEFIs was observed among adolescents with rheumatic diseases 51% [95%: 32%-70%], solid tumours 33%[95%CI: 11%-56%], Down syndrome 18%[95%CI: 6%-30%], and cystic fibrosis 17%[95%CI: 5%-29%] (**Figure 4A**). Further, among adolescents, the overall proportion of combined systemic and local reactions following the second dose of BNT162b2 was 32% [95% CI: 19%-44%]. A high proportion of reactions was observed in adolescents with rheumatic diseases 40%[95%: 16%-65%] followed by those with solid tumours 25%[95%CI: 3%-45%], Down syndrome 19%[95%CI: 10%-29%], and cystic fibrosis 27%[95%CI: 17%-38%] (**Figure 4B)**.

**Figure 4:**
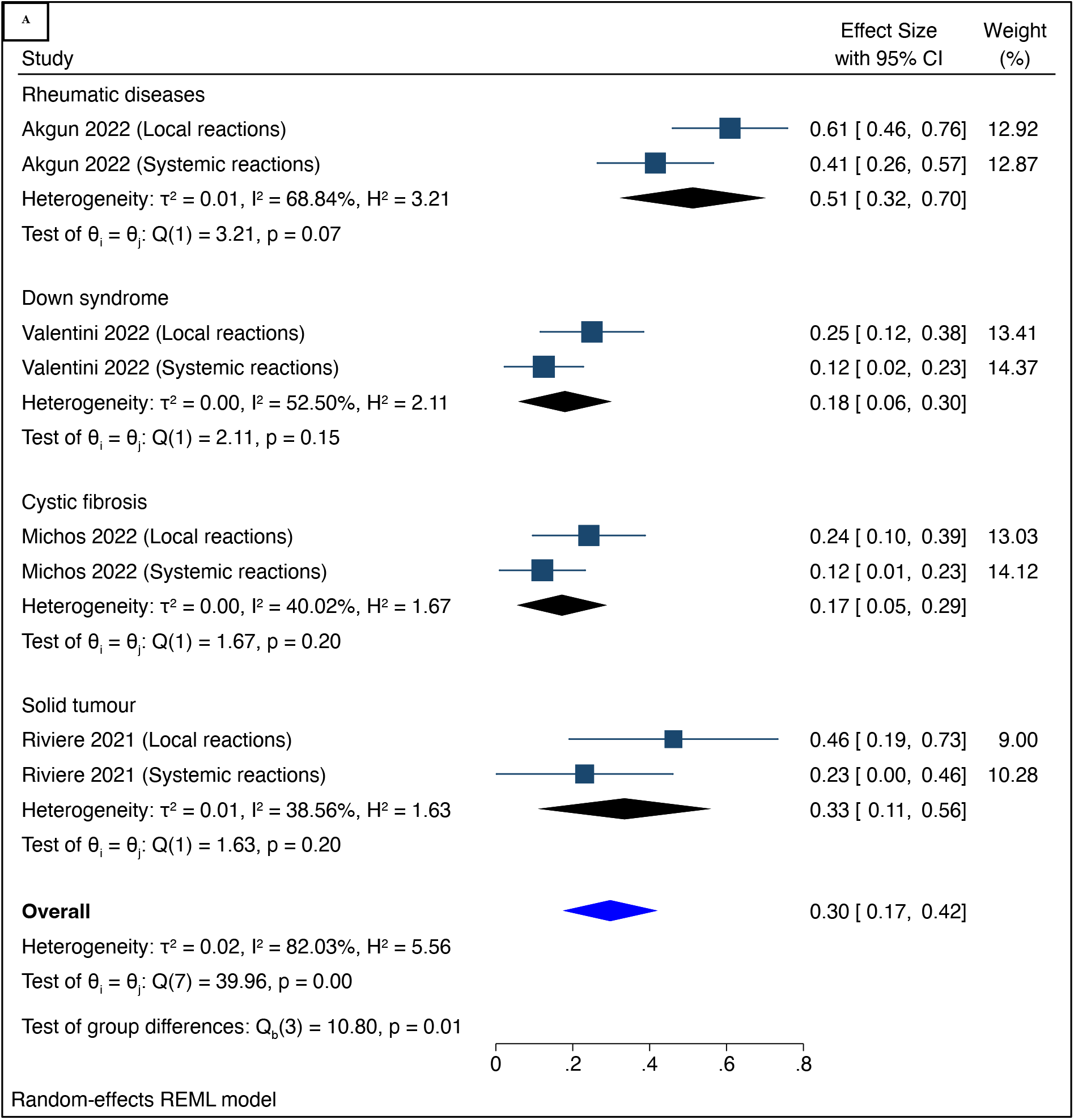
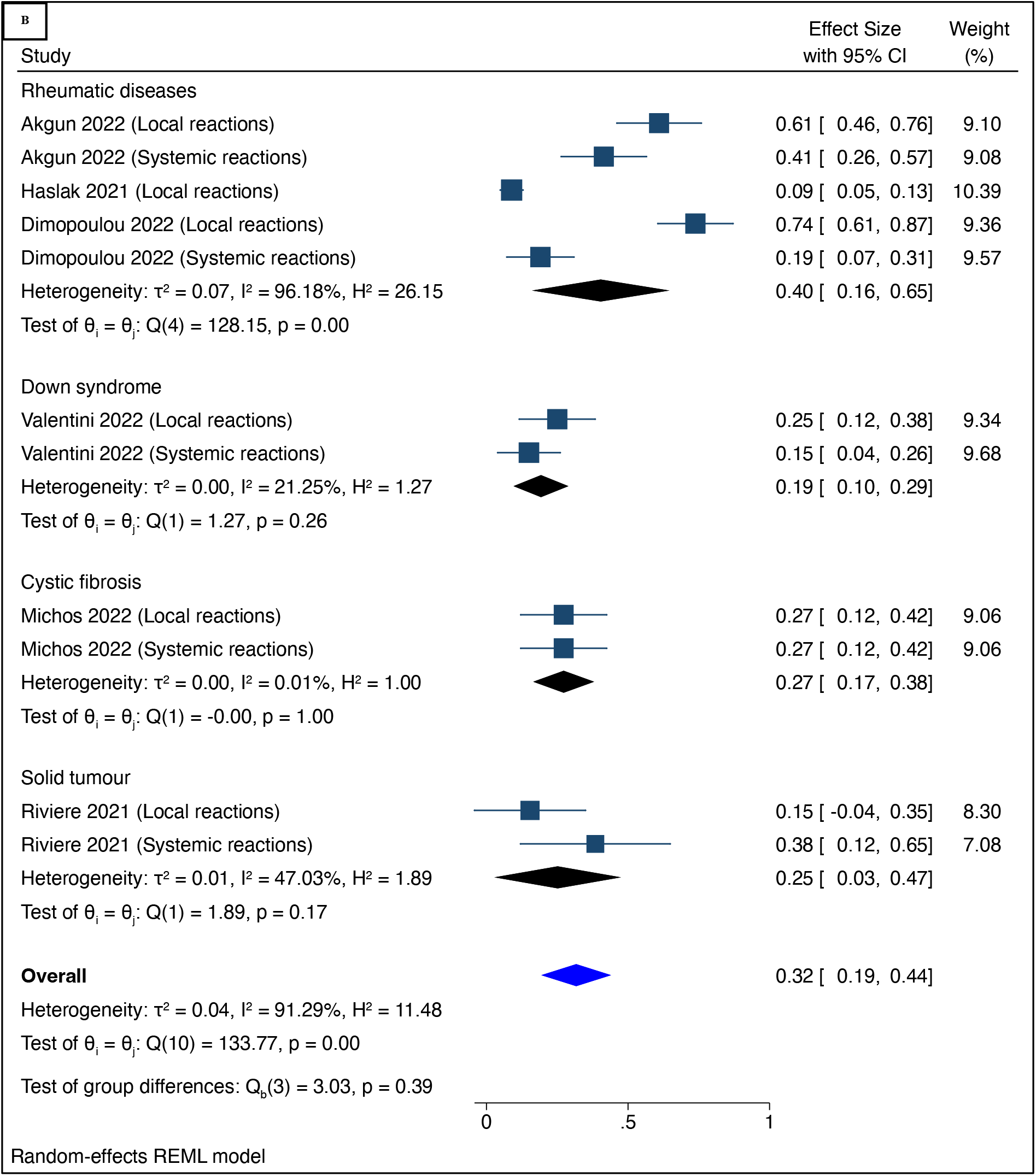
Forest plot of the proportion of combined any local and any systemic reaction among immune-compromised adolescents following receipt of the first (Panel 2A) and second (Panel 2B) doses of BNT162b2 vaccine.

### Antibody responses

The comparison of antibodies neutralization after dose 1 BNT162b2 vaccine between immunocompromised adolescents (cystic fibrosis and Down syndrome) and healthy adults was assessed in two studies. The pooled log RR revealed no statistically significant differences between the two groups, with a Log RR of 0.08 (95% CI: −0.16, 0.32, p=1, I^2^ = 0.00%) (**Figure 5A**). The comparison of antibodies neutralization after dose 2 of BNT162b2 vaccine among immunocompromised adolescents (rheumatic diseases and cystic fibrosis) and healthy adults included two studies. The pooled log RR revealed no statistically significant differences between the two groups, with a Log RR of −0.00 (95% CI: −0.24, 0.24, p=0.52, I^2^=0.00%) (**Figure 5B**).

**Figure 5:**
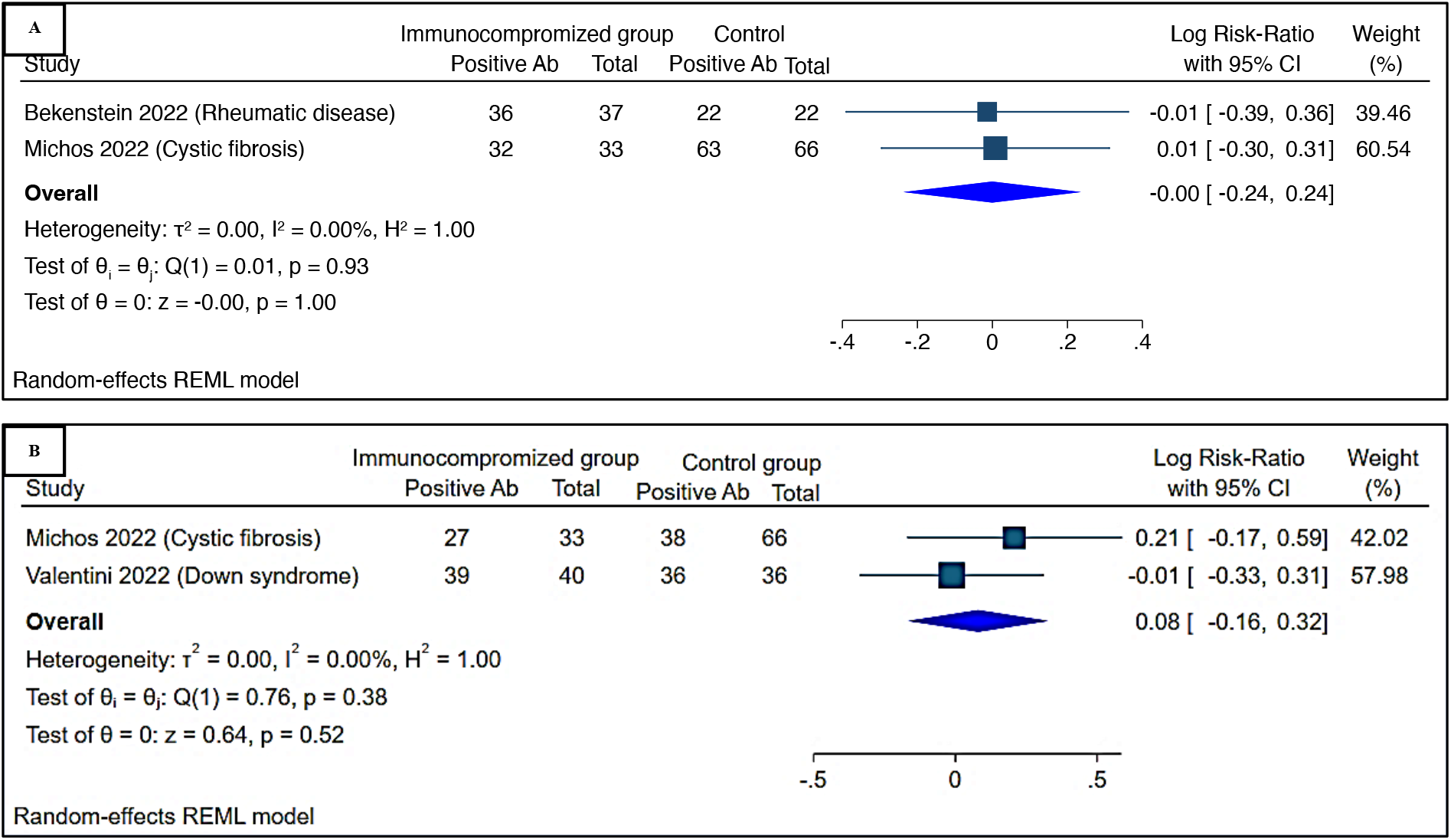
Forest plot of antibodies neutralization among immune-compromised vs healthy adolescents following receipt of the first (Panel 2A) and second (Panel 2B) doses of BNT162b2 vaccine.

The effectiveness of BNT162b2 vaccine after dose 1 in quantifying IgG (AU/ml) was reported in two studies. Valentini et al.[10] measured IgG levels in adolescents known with Down syndrome and in healthy adolescents after 21, 28, and 180 days, with mean differences (MD) IgG (AU/ml) of 146.69 (95% CI: 68.59 to 224.79), −36.99 (95% CI: −59.13 to −14.85), and − 1430.97 (95% CI: −1976.65 to −885.29). MD IgG was −405.24 (95% CI: −1345.59 to 535.12) after 21 and 180 days. Michos et al.[24] found an MD IgG of 187.14 (95% CI: −94.10 to 468.38) among cystic fibrosis vs healthy adolescents. While healthy adolescents had higher IgG − 248.25 (95% CI: −954.51 to 458.01, p=0.24, I^2^ = 99.53%), the overall pooled MD IgG was not statistically different between the two groups (**Figure 6A**). For the second dose, Valentini et al.[10] measured IgG levels in Down syndrome and healthy adolescents after 28, 45, and 180 days, with mean differences (MD) IgG (AU/ml) of −57.62(95%CI:-115.91 to 068), 0.00(95%CI: −38.16 to 38.16), and −692.74(95%CI: −719.63 to −685.85), respectively. MD IgG was −250.53 (95% CI: −686.01 to 184.96) over 28 and 180 days. In comparison, Heshin-Bekenstein et al.[4] reported a MD IgG of −56.08 (95% CI: −79.18 to −32.98) among rheumatic diseases vs healthy adolescents. Even though healthy adolescents had higher, the overall pooled MD IgG was not statistically different between the two groups, IgG −201.91 (95% CI: −524.28 to 120.45, p=0.38, I^2^ = 99.74%) (**Figure 6B**).

**Figure 6:**
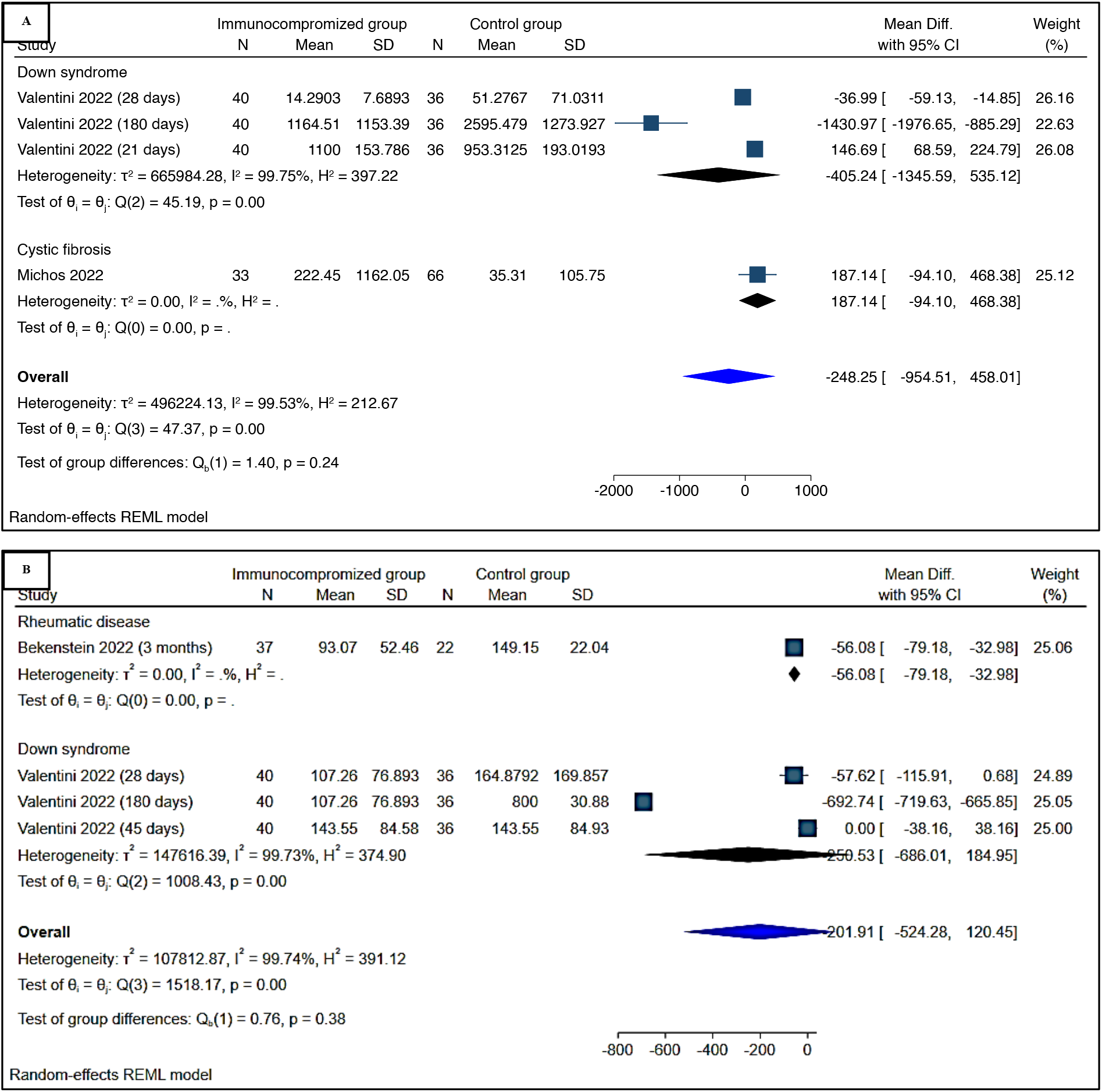
Forest plot of IgG (AU/ml) among immune-compromised vs healthy adolescents following receipt of the first (Panel 2A) and second (Panel 2B) doses of BNT162b2 vaccine.

### Publication bias

We assessed publication bias using regression tests with a random-effects model for included studies. As we performed statistical tests, publication bias was less likely for any local reaction after the second dose of BNT162b2, and no evidence of publication bias was observed for Egger’ s regression analysis (P = 0.02). Begg’ s rank correlation analysis, on the other hand, yielded a p-value of 0.003. In contrast, publication bias was clearly present for any systemic reaction following the second dose of BNT162b2 (P<0.001) for Egger’ s regression analysis and p<0.001 for Begg’ s rank correlation analysis).

## Discussion

In this systematic review and meta-analysis of mostly post-marketing studies, we found that the humoral response to BNT162b2 vaccine in AYA with various comorbidities resulting in a compromised immune system was effective after the second dose with an acceptable safety profile and without exacerbating the disease condition or causing severe adverse reactions. Many studies have reported that AYA with rheumatic diseases, severe neurological disorders, Down syndrome, type 1 diabetes, and solid tumours who received first doses of the vaccination reported mostly local reactions; the number of participants who developed AEFIs increased after the second dose. These findings are consistent with those of a randomised trial in Japanese people, in which systemic events rose from 1% to 4% after the second dose[25]. Instead, some studies found conflicting results regarding neurological complications such as Guillain-Barre syndrome and Bell’ s palsy related with the ChAdOx1 and CoronVac vaccinations but not the BNT162b2 vaccine[26].

Specificity associated with each ailment may be communicated to health care providers and caregivers. While local AEFIs were prevalent, AYA with diabetes mellitus had a higher proportion of reports of local response. Diabetes mellitus has been linked to a wide range of dermatologic disorders, many of which improve with glycaemic control[27]. As a result, it is critical to maintain appropriate diabetes management prior to delivering BNT162b2 vaccinations in order to limit the risk of morbidity. Systemic responses, such as fever, were also more common in those with diabetes, probably for the same reasons mentioned above. However, no major severe adverse events requiring hospitalization were reported, such as diabetic ketoacidosis or severe hypoglycaemia. Furthermore, modest adverse effects similar to those described in the general population were observed in adolescents with type 1 diabetes mellitus who were on different medications and monitoring systems.

Surprisingly, AYA with Down syndrome exhibited the lowest proportion of systemic responses, although having mild to moderate T cell and B cell lymphopenia and concomitant T cell abnormalities. This suggests that children and adolescents with Down syndrome have an excellent safety and tolerability profile. Another study found that when AYA with juvenile idiopathic arthritis (JIA) were given tumour necrosis factor inhibitors, the adverse responses were more common after the second dose than the first. However, no illness exacerbations occurred post vaccination[22]. Another study has reported that the vaccine had similar safety profiles, with minimal side effects occurring at a similar frequency in both adolescents with juvenile-onset autoimmune inflammatory rheumatic disorders (AIIDRs) and controls[4]. Systemic symptoms such as fever after the first dosage, as well as weariness, myalgia, and arthralgia after the second dose, were more common in adolescents with AIIDRs than in the control group, but the difference was not statistically significant. Furthermore, small increases in disease deterioration were observed, but following the first and second doses, 94.4% and 98.8% of patients maintained stable disease activity with no worsening, respectively. As instance, our findings suggest that patients with AIIRDs have acceptable safety profiles.

When antibody responses were compared after vaccination doses, there was an adequate IgG response mounted by the immune systems of AYA with the immunocompromised diseases under consideration, according to studies reporting antibody neutralization after first and second doses of vaccine. It should be highlighted that patients who tested positive for SARS-CoV-2 before to vaccination had higher IgG titers than those who tested negative (P=0.20), implying that patients who had previously been exposed to COVID-19 acquire more persistent and strong antibodies than naive patients. Considering that reinfection leading to natural immunity can be dangerous for AYA with comorbidities, it is fortunate that the second vaccine dosage exhibited comparable outcomes in increasing anti-DBD IgG titers in children receiving either conventional disease-modifying anti-rheumatic medications (cDMARDs) or biological DMARDs. Furthermore, no association was found between gender and vaccine reaction. However, treatment patterning to comorbidities affected immunologic responses to the vaccine in some cases, with lower immunologic responses observed in adolescents receiving combined therapy (conventional and biological therapy); this difference was statistically significant when compared to the biological therapy group. Despite their low levels, these youngsters were able to obtain immunizations without halting their treatment.

Some conditions not included in this meta-analysis are also of particular attention to inform decision making. For example, older adolescents with a median age of 19 years receiving an immunosuppressive regimen for a kidney transplant had poorer immunogenicity responses, with only 52% of patients showing the presence of spike antibodies after the second dose of the BNT162b2 mRNA at 4 to 8 weeks (median of 45 days), whereas younger patients in the study tended to present with better immunogenicity responses[28]. Patients on Mycophenolate Mofetil (MMF), a medication that inhibits antibody production, had the lowest immunogenicity responses. Clarkson et al.[28] reported equally modest immunogenicity responses in his cohort of patients who were also on MMF. A Canadian study looked at allergic reactions to PEG-asparaginase (PEG-ASNase), a component of BNT162b2, in 32 patients aged 12 and above who had a history of grade 2-4 allergy to PEG-ASNase, according to the Common Terminology Criteria for Adverse Events (CTCAE)[7]. However, no adverse reaction was seen 30 minutes following delivery of the vaccine. Data on adolescents with other comorbidities, particularly those prevalent in low-income countries, such as sickle cell anaemia, malnutrition, and HIV infection, is scarce. HIV is known to cause immunological dysregulation in adults, children, and adolescents and has the highest prevalence in Sub-Saharan Africa2023/01/20 10:56:00

It should be highlighted that the majority of included studies had an unclear overall risk of bias, while the rest had a high overall risk. As a result, the whole evidence base covered in this assessment has methodological limitations. While it is true that many of these studies were carried out during the height of the pandemic, when extraordinary and often emergency-response circumstances prevailed, future studies should aim to address or mitigate these limitations. This could be accomplished by addressing several difficulties. To begin, studies with comparative results (two or more arms) should be carefully adjusted to account for potential confounders, while also making an effort to recruit a comparable control group to reduce the chance of false findings. Such comparison evidence is desirable, but if a single-arm study is required, the data should be stratified by potential effect modifiers and confounders to investigate their impact on the outcomes. Second, detailed reporting of participant selection as well as if - and how - vaccination status was confirmed is necessary, as confusion in these domains is frequently caused by a lack of reported data. This data should describe the inclusion of a diverse group of patients who match the eligibility requirements; convenience selection should be avoided to reduce the possibility of volunteer bias. Furthermore, there must be explicit reporting of the measures used and sources consulted to ensure vaccination status. Third, attrition should be reduced to the greatest extent practicable by follow-up and assuring the availability of data-generating resources. Where attrition is unavoidable, it should be clearly recorded as individuals lost to follow-up or with missing data from the initial eligible sample, along with reasons, and suitable statistical procedures should be employed to account for missing data where substantial attrition is encountered. Exclusions based on the investigator should be kept to a minimum as well. Fourth, the determination of outcomes, particularly safety outcomes, should be standardised and objectively assessed. This is especially significant in comparative studies, because subjective participant-reported metrics may lead to erroneous nocebo effects. Fifth, any future studies should have clear prospective *a priori* protocols or statistical analysis plans in place to allow for an assessment of the risk of bias associated with probable selective outcomes presented. While it is recognized that the relative rarity of the conditions studied in the review may result in small sample sizes, this should be avoided to the greatest extent possible by including larger groups of potentially eligible participants to improve generalisability and reduce the risk of fragile or chance findings.

## Conclusion

AYA with rheumatic disorders, severe neurodisabilities, Down syndrome, type 1 diabetes, and solid tumours who received the COVID-19 BNT162b2 mRNA vaccine showed good immunogenicity and safety. While illness specificity should be recognized to guide practice, there is an urgent need to improve the design of post-marketing studies of AYA with comorbidity, as well as research on comorbidities that are prevalent in low-resource settings, such as HIV infection, to ensure vaccine equality.

## Data Availability

All data produced in the present work are contained in the manuscript

## Contributors

PK, CSW and GG conceived the study. PK, LB and MM did the literature search. PK and LB did the study selection. JT, LB, AB, and PK extracted the relevant information. PK accessed and verified the data. AB supervised the risk of study section. PK and JT synthesized the data. PK, MM, AA, LB and AB wrote the first draft. CSW and GG supervised the overall work. All authors critically revised successive drafts of the paper. All authors had full access to all the data in the study, read, and approved the final manuscript, and had final responsibility for the decision to submit for publication

## Declaration of interests

All authors declare no competing interests.

## Data sharing

All data generated or analysed during this study are included in this published Article and the appendix.

## Acknowledgments

We acknowledge the South African Medical Research Council for supporting the work of some of the authors and for providing funding for open access publication of this study.

## Notes

### Competing Interest Statement

The authors have declared no competing interest.

