## Supplementary material for "Immunogenicity, Safety and Effectiveness of COVID-19 Pfizer-BioNTech (BNT162b2) mRNA Vaccination in Immunocompromised Adolescents and Young Adults: A systematic Review and Meta-Analyses": NA

### Supplementary materials

#### Method and materials (continued)

##### Safety description

Overall, outcomes related to safety were assessed at any time point and included: any AEFIs, any adverse events (AEs); serious, non-serious and leading to discontinuation, any adverse events of special interest (AESIs), local injection site reactions, local reactions (e.g., erythema, pain or swelling), systemic reactions (e.g. myalgia, rash, abdominal pain, itching, arthralgia, muscle pain, chills, fatigue, fever, headache, nausea, vomiting or diarrhea), any related events, any life-threatening related events, need for medical attention due to adverse reaction (e.g. antipyretic or analgesic treatment, hospitalization, inability to perform daily activities or attend school/work, telehealth consults, attendance to clinic or emergency room or taking medication for side effects), serious adverse events (SAEs) (e.g. chest pain, tachycardia, dyspnea, myocarditis, myopericarditis or severe allergic reactions), non-serious AEs, AEs leading to discontinuation from the study, AEFI by system organ class (SOC) (e.g. infections and infestations; general disorders and administration site conditions; nervous system disorders; sleeping disturbancee; musculoskeletal and connective tissue disorders; respiratory, thoracic and mediastinal disorders; cardiac disorders; arrhythmias; injury, poisoning and procedural complications; vascular disorders; haemorrhagic diseases; thromboembolism; reproductive system and breast disorders; hepato-renal syndrome; gastrointestinal disorders or psychiatric disorders), unsolicited or unbalanced AEFIs (e.g. autoimmune diseases, Guillain-Barré syndrome; acute aseptic arthritis or Bell's palsy), hypersensitivity, lymphadenopathy and death. Then, we classified all adverse events as local and systemic AEFI, serious AEFI, AESI, health impact, and unsolicited AEFI by SOC following any dose of BNT162b2 in adolescents

##### Data extraction and management

Study characteristics and outcomes data were extracted by a single reviewer and crosschecked by a second reviewer to ensure consistency. Extractions were conducted using a pre-piloted extraction form set up in Covidence, which included the following fields: (1) identification of the study: country in which the study was conducted and time period over which the study was conducted, (2) methods: aim of the study and study design, participants-mean age and age group of participants included in the study, total number of participants included, number of participants per group (if reported), (3) interventions: follow-up period after vaccine administration (first and second dose), primary doses or booster administered and (4) safety outcomes: type of AEFI and number of events in each group and sub-age-group as well. (5) neutralising antibodies (type, titter and time) and (6) Level of efficacy of effectiveness (proportion and period of time after vaccination). At any point of time, discrepancies were resolved through consensus or adjudication by a senior review author. A Characteristics of included studies table was completed using descriptive information extracted from studies and exported into excel sheet from Covidence.

#### Supplementary tables and box

**Supplementary Table 1: Definition of Safety Outcome (adapted from Wu et al)**

| Outcome | Definition |
|---|---|
| Adverse event following immunization (AEFI) | An AEFI is any untoward medical occurrence which follows immunization and which does not necessarily have a causal relationship with the usage of the vaccine. The adverse event may be any unfavourable or unintended sign, abnormal laboratory finding, symptom or disease. |
| Serious adverse events (SAE) | A serious adverse event is any untoward medical occurrence that at any dose results in death, requires inpatient hospitalization or prolongation of existing hospitalization, results in persistent or significant disability/incapacity, or is life-threatening. |
| Solicited adverse events | Solicited adverse events include prospectively self-collected occurrences of local and systemic reactions. Participants were usually asked to monitor and record local reactions, systemic events, and antipyretic medication usage for 7 days following each administration. |
| Local reactions | Local reactions included pain at the injection site, redness, swelling, induration, etc. |
| Systemic reactions | Systemic reactions included headache, myalgia, fever, fatigue, vomiting, diarrhea, muscle pain, etc. |
| Unsolicited adverse events | Unsolicited AEs would be represented in the AE domain unless they were classified as solicited adverse event. |
| Adverse event of special interest (AESI) | An adverse event of special interest (serious or non-serious) is one of scientific and medical concern specific to vaccine, for which ongoing monitoring can be appropriate. |
| Withdrawal due to adverse events | The number of participants reported as withdrawn from clinical trial due to adverse events whether related to study intervention or not. |
| Death | The number of participants reported for death regardless of causality. |

### Supplementary Table 2: Characteristic of included studies

| Study ID, settings, design and average age of participants | Condition | Objective | Dose type | Outcomes of Interest | Event in immunocompromised | Total in immunocompromised | Event Control | Total Control | Time | Mean (SE) in AU/ml/Immunocompromised | Total Immunocompromised | Mean (SE) in Au/ml Control | Total Control | Authors conclusion |
|---|---|---|---|---|---|---|---|---|---|---|---|---|---|---|
| Akgun et al., 2022/Turkey, Prospective cohort of 15.4 ± 1.5 years | Rheumatic diseases | To examine vaccine antibody response of children and adolescents with rheumatic diseases in relation to confounding factors. To evaluate systemic and local side effects of the BNT162b2 mRNA vaccine in children with paediatric rheumatic diseases. | Dose 1 | Pain | 24 | 41 | | | | | | | | Paediatric rheumatic diseases patients receiving immunomodulatory treatments were able to mount an effective humoral response after two dose regimens of BNT162b2 mRNA vaccine safely without interrupting their current treatments. |
| | | | | Swelling | 2 | 41 | | | | | | | | |
| | | | | Erythema | 1 | 41 | | | | | | | | |
| | | | | Fever | 5 | 41 | | | | | | | | |
| | | | | Muscles aches | 6 | 41 | | | | | | | | |
| | | | | Headaches | 10 | 41 | | | | | | | | |
| | | | | Fatigue | 13 | 41 | | | | | | | | |
| | | | Dose 2 | Pain | 20 | 41 | | | | | | | | |
| | | | | Swelling | 2 | 41 | | | | | | | | |

| | | | | | | | | | | | | | | |
|---|---|---|---|---|---|---|---|---|---|---|---|---|---|---|
| | | | | Erythema | 1 | 41 | | | | | | | | |
| | | | | Fever | 6 | 41 | | | | | | | | |
| | | | | Muscles aches | 7 | 41 | | | | | | | | |
| | | | | Headaches | 11 | 41 | | | | | | | | |
| | | | | Fatigue | 12 | 41 | | | | | | | | |
| Bekenstein et al., 2022/Israel and Slovenia, Prospective cohort study, adolescents ( 12– | Rheumatic diseases | To explore safety and immunogenicity of the mRNA COVID-19 vaccine among | Dose 1 | Pain | 64 | 88 | | | | | | | | Evidence of good short-term vaccine safety and adequate humoral immune |

| | | | | | | | | | | | | | | |
|---|---|---|---|---|---|---|---|---|---|---|---|---|---|---|
| 18 years) and young adults (18–21 years) | | adolescents with juvenile-onset AIIRDs treated with immunomodulatory medications and compared it with the results among healthy children | | Swelling | 8 | 88 | | | | | | | | response in the unique population of adolescents with juvenile-onset AIIRDs. |
| | | | | Erythema | 2 | 88 | | | | | | | | |
| | | | | Itching | 3 | 88 | | | | | | | | |
| | | | | Pruritus | 1 | 88 | | | | | | | | |
| | | | | Fever | 14 | 88 | | | | | | | | |
| | | | | Vomiting | 2 | 88 | | | | | | | | |
| | | | | Nausea | 6 | 88 | | | | | | | | |
| | | | | Running nose | 4 | 88 | | | | | | | | |
| | | | | Muscles aches | 19 | 88 | | | | | | | | |
| | | | | Joint pain | 10 | 88 | | | | | | | | |
| | | | | Chills | 11 | 88 | | | | | | | | |
| | | | | Headache | 22 | 88 | | | | | | | | |
| | | | | Feeling unwell | 24 | 88 | | | | | | | | |
| | | | | Weakness | 25 | 88 | | | | | | | | |
| | | | | Fatigue | 29 | 88 | | | | | | | | |
| | | | | Exacerbation | 1 | 88 | | | | | | | | |
| | | | Dose 2 | Antibody seropositivity | 36 | 37 | 22 | 22 | 3 months | | | | | |
| | | | | IgG dosage | | | | | 3 months | 93.07 (52.46) | 37 | 149.15 (22.04) | 22 | |
| Dimopoulou et al., 2022/Greece, Single-center, cohort study of 17 (16–21) years | Rheumatic diseases | To evaluate safety and tolerability of BNT162b2 COVID-19 vaccine in adolescents | Dose 2 | Pain | 21 | 42 | | | | | | | | mRNA vaccines appear to be safe and well tolerated in adolescents with JIA receiving treatment with TNF inhibitors |
| | | | | Swelling | 12 | 42 | | | | | | | | |
| | | | | Erythema | 31 | 42 | | | | | | | | |

| | | | | | | | | | | | | | | |
|---|---|---|---|---|---|---|---|---|---|---|---|---|---|---|
| | | | | Muscles aches | 5 | 42 | | | | | | | | |
| | | | | Joint pain | 5 | 42 | | | | | | | | |
| | | | | Headache | 7 | 42 | | | | | | | | |
| | | | | Fatigue | 6 | 42 | | | | | | | | |
| Haslak et al., 2021/Turkey, Cross-sectional study, of 15.34 (12.02-20.92) | Rheumatic diseases | To examine vaccine related adverse events of BNT162b2 messenger COVID-19 vaccine | Dose 1 | Fever | 14 | 191 | | | | | | | | Acceptable safety profile of COVID-19 vaccines and encouragement of children with IRD to be vaccinated. |
| | | | | Vomiting | 23 | 191 | | | | | | | | |
| | | | | Nausea | 2 | 191 | | | | | | | | |
| | | | | Headache | 10 | 191 | | | | | | | | |
| | | | | Fatigue | 25 | 191 | | | | | | | | |
| | | | Dose 2 | Fever | 12 | 191 | | | | | | | | |
| | | | | Vomiting | 6 | 191 | | | | | | | | |
| | | | | Headache | 14 | 191 | | | | | | | | |
| | | | | Fatigue | 19 | 191 | | | | | | | | |
| King et al., 2022/UK, Prospective cohort of 12-15 years | Neurodisa bilities | To inform the risk–benefit for subsequent COVID-19 vaccinations | Dose 1 | Erythema | 1 | 26 | | | | | | | | Mild/moderate adverse reactions except for one child |
| | | | | Fever | 3 | 26 | | | | | | | | |
| | | | | Feeling unwell | 10 | 26 | | | | | | | | |
| | | | | Fatigue | 19 | 26 | | | | | | | | |
| | | | Dose 2 | Erythema | 2 | 22 | | | | | | | | |
| | | | | Fever | 3 | 22 | | | | | | | | |
| | | | | Feeling unwell | 3 | 22 | | | | | | | | |
| | | | | Fatigue | 6 | 22 | | | | | | | | |

| | | | | | | | | | | | | | | |
|---|---|---|---|---|---|---|---|---|---|---|---|---|---|---|
| Valentini et al., 2022/Italy, Prospective cohort of 17.90 (±4.59) | Down syndrome | To evaluate the safety of mRNA vaccination in individuals with DS. To measured SARS-CoV-2 specific antibodies over time. To correlate humoral immune response of individuals with DS with those of the healthy controls (HC) | Dose 1 | Pain | 10 | 40 | | | | | | | | Individuals with DS exhibit a good humoral response to the BNT162b2 vaccine; however, similarly to in HC, the immune response wanes over time. |
| | | | | Erythema | 1 | 40 | | | | | | | | |
| | | | | Fever | 2 | 40 | | | | | | | | |
| | | | | Muscles aches | 1 | 40 | | | | | | | | |
| | | | | Chills | 1 | 40 | | | | | | | | |
| | | | | Headache | 1 | 40 | | | | | | | | |
| | | | | Fatigue | 1 | 40 | | | | | | | | |
| | | | | Antibody seropositivity | 39 | 40 | 36 | 36 | | | | | | |
| | | | Dose 2 | Pain | 10 | | | | | | | | | |
| | | | | Erythema | 2 | | | | | | | | | |
| | | | | Fever | 4 | | | | | | | | | |
| | | | | Muscles aches | 1 | | | | | | | | | |
| | | | | Headache | 1 | | | | | | | | | |
| | | | | Fatigue | 2 | | | | | | | | | |
| | | | Dose 1 | IgG dosage | | | | | 21 days | 14.29(7.68) | 40 | 51.27(71.03) | 36 | |
| | | | | | | | | | 28 days | 1164.51(1153.39) | 40 | 2595.47(1273.92) | 36 | |
| | | | | | | | | | 180 days | 1100(153.78) | 40 | 953.31(193.01) | 36 | |
| | | | Dose 2 | IgG dosage | | | | | 21 days | 107.26(76.89) | 40 | 164.87(169.85) | 36 | |
| | | | | | | | | | 28 days | 107.26(76.89) | 40 | 800(30.88) | 36 | |
| | | | | | | | | | 180 days | 143.55(84.58) | 40 | 143.55(84.93) | 36 | |

| | | | | | | | | | | | | | | |
|---|---|---|---|---|---|---|---|---|---|---|---|---|---|---|
| Michos et al., 2022/Italy, Prospective cohort of 19.6 (17.6–24.3) | Cystic fibrosis | To investigate immunogenicity of SARS-CoV-2 BNT162b2 vaccine and its association with epidemiological and clinical parameters in a cohort of CF patients and to compare it with a cohort of healthy individuals | Dose 1 | Antibody seropositivity | 27 | 33 | 38 | 66 | | | | | | BNT162b2 vaccine appears to be immunogenic with limited adverse events in CF population. |
| | | | Dose 2 | Antibody seropositivity | 32 | 33 | 63 | 66 | 1 month | | | | | |
| | | | Dose 1 | IgG dosage | | | | | | 222.45(1162.05) | 33 | 35.31(105.75) | 66 | |
| Piccini et al., 2022/Italy, Retrospective study, 18.4 ± 2.4 | Type 1 diabetes | To evaluate adverse effects, possible glycemic control modification and temporary insulin dose adjustment in youth with T1D, users of different levels of technology, who completed a whole COVID-19 vaccination cycle | Dose 1 | Pain | 28 | 39 | | | | | | | | Vaccination not associated to significant perturbation of glycemic control in adolescents and young adults with T1D, and, if elevation of glucose values occurs, it is mild, transient, tolerable and not requiring insulin dose adjustment. Side effects are mild and similar to those reported in the general population. All in all, this information could be of clinical use when counseling families in order to reassure them about SARS-CoV-2 vaccination in youth with T1D. |
| | | | | Fever | 5 | 39 | | | | | | | | |
| | | | | Muscles aches | 5 | 39 | | | | | | | | |
| | | | | Headache | 7 | 39 | | | | | | | | |
| | | | | Weakness | 16 | 39 | | | | | | | | |
| | | | Dose 2 | Pain | 25 | 39 | | | | | | | | |
| | | | | Fever | 10 | | | | | | | | | |
| | | | | Muscles aches | 5 | 39 | | | | | | | | |
| | | | | Headache | 12 | 39 | | | | | | | | |
| | | | | Weakness | 17 | 39 | | | | | | | | |
| Riviere et al., 2021/France, Retrospective | Solid tumour | To evaluate safety and efficacy of BNT162b2 vaccine in | Dose 1 | Pain | 6 | 13 | | | | | | | | Preliminary experience with RNA vaccines in AYA with |

| | | | | | | | | | | | | | | |
|---|---|---|---|---|---|---|---|---|---|---|---|---|---|---|
| study, Median: 17 years | | adolescents and young adults (AYA) with solid tumour. | | Fever | 2 | 13 | | | | | | | | solid tumours and report a good safety profile and excellent immunogenicity |
| | | | | Headache | 1 | 13 | | | | | | | | |
| | | | | Fatigue | 2 | 13 | | | | | | | | |
| | | | Dose 2 | Pain | 2 | 13 | | | | | | | | |
| | | | | Fever | 4 | 13 | | | | | | | | |
| | | | | Vomiting | 1 | 13 | | | | | | | | |
| | | | | Fatigue | 5 | 13 | | | | | | | | |

### Supplementary Table 3: Risk of bias in included studies (ROBINS-I)

| **Domain** | | **Akgün 2022 (cohort)** | **Dimopoulou 2022 (cohort)** | **Haslak 2022 (cross-sectional)** | **Heshin-Bekenstein 2022 (controlled cohort)** | **King 2022 (cohort/active surveillance)** | **Michos 2022 (controlled cohort)** | **Piccini 2022 (retrospective cohort)** | **Riviere 2021 (retrospective cohort)** | **Valentini 2022 (controlled cohort)** |
|---|---|---|---|---|---|---|---|---|---|---|
| **Bias due to confounding** | **Judgment** | Unclear | Low | Low | High | Unclear | High | Unclear | Unclear | High |
| | **Supporting text** | Efficacy results are stratified by diagnoses and treatments. Safety results do not appear to be stratified. | Safety results are analysed by type of diagnosis and treatments. | Safety results are analysed by type of diagnosis, treatments, and other demographic characteristics. | Efficacy results are stratified by treatments, no adjustment for confounders reported, however, age-matching was identified by authors as sub-optimal since controls were younger than participants with rheumatic disease. Safety results do not appear to be stratified or adjusted for confounders. | Safety results are not stratified. | Efficacy results are analysed by demographic and clinical characteristics, no adjustment for confounders reported. A lack of age and gender matching was reported by the authors. Safety results do not appear to be stratified or adjusted. | Safety results are not stratified. | Efficacy and safety results are not stratified. | Efficacy results are analysed by demographic and clinical characteristics, no adjustment for confounders reported. Authors identified the lack of age-matched controls as a limitation. Safety results do not appear to be stratified or adjusted. |
| **Bias in selection of participants into the study** | **Judgment** | Unclear | Unclear | Unclear | Unclear | Unclear | Unclear | Unclear | Unclear | Unclear |
| | **Supporting text** | Patients were invited by phone call – it is unclear | Insufficient information on | Data were captured by web-based | Insufficient information on recruitment of | Clinician-identified patients were invited | Insufficient information on recruitment of | Insufficient information on inclusion into | Insufficient information on recruitment. | Insufficient information on recruitment of |

| Domain | | Akgün 2022 (cohort) | Dimopoulou 2022 (cohort) | Haslak 2022 (cross-sectional) | Heshin-Bekenstein 2022 (controlled cohort) | King 2022 (cohort/active surveillance) | Michos 2022 (controlled cohort) | Piccini 2022 (retrospective cohort) | Riviere 2021 (retrospective cohort) | Valentini 2022 (controlled cohort) |
|---|---|---|---|---|---|---|---|---|---|---|
| | | whether volunteer bias may be present. | recruitment. Informed consent was sought, volunteer bias could not be ruled out. | survey, with no information on recruitment – it is unclear whether volunteer bias may be present. | patients with rheumatic disease as well as healthy controls. These groups were recruited from different centres. Informed consent was sought. | following informed medical consent. It is unclear whether volunteer bias may be present. | patients with cystic fibrosis as well as healthy controls. These groups were systematically different as controls were healthcare workers. Informed consent was sought. | this retrospective study. | Informed consent was sought, volunteer bias could not be ruled out. | patients with Down syndrome as well as healthy controls. These groups were systematically different as controls were healthcare workers. Informed consent was sought. |
| **Bias in classification of interventions** | **Judgment** | Low | Low | Unclear | Low | Low | Unclear | Unclear | Low | Unclear |
| | **Supporting text** | Patients were vaccinated with the BNT162b2 mRNA vaccine post-recruitment. | Patients were vaccinated with the BNT162b2 mRNA vaccine post-recruitment. | Vaccination status was verified by phone calls and national registries, however, two types of COVID-19 vaccine was available. It is not clear how the type of vaccine received was determined. | Patients were vaccinated with the BNT162b2 mRNA vaccine post-recruitment. | Patients were vaccinated with the BNT162b2 mRNA vaccine post-recruitment. | Authors report that both groups were vaccinated with the BNT162b2 mRNA vaccine. It is unclear if this was post-recruitment, or whether vaccination status was confirmed. | It is not reported how vaccination status was verified. | Patients were vaccinated with the BNT162b2 mRNA vaccine post-recruitment. | Authors report that both groups were vaccinated with the BNT162b2 mRNA vaccine. It is unclear if this was post-recruitment, or whether vaccination status was confirmed. |
| **Bias due to deviations from intended interventions** | **Judgment** | Unclear | Unclear | Low | Unclear | Unclear | Unclear | Unclear | Unclear | Unclear |
| | **Supporting text** | It is not reported whether – or how - vaccination status of participants was determined. | It is not reported how vaccination status of participants was determined. | Vaccination status was verified by phone calls and national registries. | It is not reported whether – or how - vaccination status of participants was determined. | It is not reported whether – or how - vaccination status of participants was determined. | Authors report that both groups were vaccinated with the BNT162b2 mRNA vaccine. It is unclear if this was post-recruitment, or whether vaccination status was confirmed. | It is not reported how vaccination status of participants was determined. | It is not reported whether – or how - vaccination status of participants was determined. | Authors report that both groups were vaccinated with the BNT162b2 mRNA vaccine. It is unclear if this was post-recruitment, or whether vaccination status was confirmed. |
| **Bias due to missing data** | **Judgment** | Unclear | Unclear | Low | High | Unclear | Unclear | Unclear | Low | Low |
| | **Supporting text** | No withdrawals or loss to follow up is reported, but some investigator-driven exclusions, e.g. exclusion of patients without complete | No details on how many participants were potentially eligible, therefore attrition could not be assessed. | A total of 39/285 potentially eligible patients (14%) were excluded, but with reasons that were considered reasonable. | Approximately 3% loss to follow-up in the group with rheumatic disease following the second dose, with no loss to follow-up in the control group. High attrition for/low availability of | No details on how many participants were potentially eligible, therefore attrition could not be assessed. | No withdrawals or loss to follow up is reported, but some investigator-driven exclusions are implied, e.g. exclusion of patients without full vaccination. This does not align with | No withdrawals or loss to follow up is reported, but some investigator-driven exclusions are implied, e.g. inclusion of only patients | Approximately 23% loss to follow-up, but for valid reasons, and no investigator-driven exclusions are reported. | No withdrawals or loss to follow up is reported and investigator-driven exclusion of individuals with Down syndrome, for the presence of antibodies at baseline, is |

| Domain | | Akgün 2022 (cohort) | Dimopoulou 2022 (cohort) | Haslak 2022 (cross-sectional) | Heshin-Bekenstein 2022 (controlled cohort) | King 2022 (cohort/active surveillance) | Michos 2022 (controlled cohort) | Piccini 2022 (retrospective cohort) | Riviere 2021 (retrospective cohort) | Valentini 2022 (controlled cohort) |
|---|---|---|---|---|---|---|---|---|---|---|
| | | vaccination schedule (14% of total sample), do not align with the review question. | | | immunogenicity data (41% in rheumatic disease group and 55% in the control group provided data) due to limited availability of serum samples. No further exclusions reported. | | the review question, though its impact could not be numerically assessed. | who underwent a whole vaccination cycle. This does not align with the review question, though its impact could not be numerically assessed. | | considered reasonable. |
| **Bias in measurement of outcomes** | **Judgment** | Unclear | Unclear | Unclear | High | Unclear | Unclear | Unclear | Unclear | Unclear |
| | **Supporting text** | Efficacy outcomes were clearly defined and laboratory-determined, with minimal risk of ascertainment bias. Safety outcomes were participant-reported, using a structured questionnaire; while this may have introduced bias this would not be differential, given the single-arm design. | Safety outcomes were participant-recorded, using a diary card; while this may have introduced bias this would not be differential, given the single-arm design. | It is not clear whether safety was objectively assessed: '…experienced adverse events of the participants were verified by phone calls and national registries.' | Efficacy outcomes were clearly defined and laboratory-determined, with minimal risk of ascertainment bias. Safety outcomes, however, were participant-reported using a telephonic questionnaire. Given the controlled nature of the study, this may have biased measures of association. | Parents were asked to record side effects in a diary and were followed up with a telephone call; while this may have introduced bias this would not be differential, given the single-arm design. | Efficacy outcomes were clearly defined and laboratory-determined, with minimal risk of ascertainment bias. Safety outcomes were participant-reported; while this may have introduced bias this would not be differential, given the single-arm safety comparison. | Symptoms after first and second dose recorded in medical files were retrospectively investigated, either face to face or via telemedicine. | Efficacy outcomes were clearly defined and laboratory-determined, with minimal risk of ascertainment bias. Safety outcomes were participant-reported; while this may have introduced bias this would not be differential, given the single-arm design. | Efficacy outcomes were clearly defined and laboratory-determined, with minimal risk of ascertainment bias. Safety outcomes were participant-reported; while this may have introduced bias this would not be differential, given the single-arm safety comparison. |
| **Bias in selection of the reported result** | **Judgment** | Unclear | Unclear | Unclear | Unclear | Unclear | Unclear | Unclear | Unclear | Unclear |
| | **Supporting text** | We did not have access to the protocol, therefore selective reporting could not be assessed. | We did not have access to the protocol, therefore selective reporting could not be assessed. | We did not have access to the protocol, therefore selective reporting could not be assessed. | We did not have access to the protocol, therefore selective reporting could not be assessed. | We did not have access to the protocol, therefore selective reporting could not be assessed. | We did not have access to the protocol, therefore selective reporting could not be assessed. | We did not have access to the protocol, therefore selective reporting could not be assessed. | We did not have access to the protocol, therefore selective reporting could not be assessed. | We did not have access to the protocol, therefore selective reporting could not be assessed. |
| **Other source of bias** | **Judgment** | Unclear | Low | Unclear | Unclear | Low | Unclear | Low | Unclear | Unclear |
| | **Supporting text** | Lack of control group, small sample size, short | None identified. | Details on medication doses, duration of treatment and | Disease diversity limited conclusions regarding the effect of | None identified. | Limited numbers of patients with cystic fibrosis. | None identified. | Small sample size, patients were limited to those with solid | Small sample size. |

| Domain | | Akgün 2022 (cohort) | Dimopoulou 2022 (cohort) | Haslak 2022 (cross-sectional) | Heshin-Bekenstein 2022 (controlled cohort) | King 2022 (cohort/active surveillance) | Michos 2022 (controlled cohort) | Piccini 2022 (retrospective cohort) | Riviere 2021 (retrospective cohort) | Valentini 2022 (controlled cohort) |
|---|---|---|---|---|---|---|---|---|---|---|
| | | post-vaccine follow-up. | | disease activity not available. Some groups with vaccine effect-modifying characteristics were grouped together. | immunomodulatory medication and disease type on efficacy. | | | | tumours limiting generalisability to all paediatric oncology patients, short post-vaccine follow-up. | |
| **Overall bias** | **Judgment** | Unclear | Unclear | Unclear | High | Unclear | High | Unclear | Unclear | High |
| | **Supporting text** | Bias due to confounding and deviations from intended interventions as well as selection bias, attrition bias, ascertainment bias and selective outcome reporting could not be ruled out. Several other limitations were identified. | Bias due to deviations from intended interventions as well as selection bias, attrition bias, ascertainment bias and selective outcome reporting could not be ruled out. | Selection bias, classification bias, ascertainment bias, and selective outcome reporting could not be ruled out. Several other limitations were identified. | High risk of confounding, attrition bias and ascertainment bias. Bias due to deviations from intended interventions as well as selection bias and selective outcome reporting could not be ruled out. Several other limitations were identified. | Bias due to confounding and deviations from intended interventions as well as selection bias, attrition bias, ascertainment bias and selective outcome reporting could not be ruled out. | High risk of confounding. Selection bias, classification bias, deviation from intended interventions, attrition bias, ascertainment bias, and selective outcome reporting could not be ruled out. Other limitations were also identified. | Confounding, selection bias, classification bias, deviation from intended interventions, attrition bias, ascertainment bias, and selective outcome reporting could not be ruled out. | Confounding, selection bias, deviation from intended interventions, ascertainment bias, and selective outcome reporting could not be ruled out. Several other limitations were identified. | High risk of confounding. Selection bias, classification bias, deviation from intended interventions, ascertainment bias, and selective outcome reporting could not be ruled out. Other limitations were also identified. |

#### Supplementary box 1: Search terms built for MEDLINE (PubMed)

("COVID-19"[Mesh] OR "SARS-CoV-2"[Mesh] OR "COVID-19 Vaccines"[Mesh] OR "COVID-19 Serological Testing"[Mesh] OR "COVID-19 Nucleic Acid Testing"[Mesh] OR "SARS-CoV-2 variants" [Supplementary Concept] OR "COVID-19 drug treatment" [Supplementary Concept] OR "COVID-19 serotherapy" [Supplementary Concept] OR "2019-nCoV" OR "2019nCoV" OR "cov 2" OR "Covid-19" OR "sars coronavirus 2" OR "sars cov 2" OR "SARS-CoV-2" OR "severe acute respiratory syndrome coronavirus 2" OR "coronavirus 2" OR "COVID 19" OR "COVID-19" OR "2019 ncov" OR "2019nCoV" OR "corona virus disease 2019" OR "cov2" OR "COVID-19" OR "COVID19" OR "nCov 2019" OR "nCoV" OR "new corona virus" OR "new coronaviruses" OR "novel corona virus" OR "novel coronaviruses" OR "SARS Coronavirus 2" OR "SARS2" OR "SARS-COV-2" OR "Severe Acute Respiratory Syndrome Coronavirus 2")

AND

(BNT162b2[All Fields] OR Pfizer-BioNTech[All Fields] OR Pfizer/BioNTech[All Fields] OR Comirnaty[All Fields])

AND

(tolerability[Title/Abstract] OR reactogenicity[Title/Abstract] OR safe*[Title/Abstract] OR side effect[Title/Abstract] OR adverse event[Title/Abstract] OR adverse effect[Title/Abstract] OR adverse reaction[Title/Abstract] OR adverse OR efficacy[Title/Abstract] OR effectiveness[Title/Abstract] OR outcome[Title/Abstract] OR undesirable effect*[Title/Abstract] OR treatment emergent[Title/Abstract] OR toxicity[Title/Abstract])

AND

("Adolescent"[Mesh] OR "Adolescen*"[Title/Abstract] OR "Teen*"[Title/Abstract] OR "Youth*"[Title/Abstract] OR "juvenile*"[Title/Abstract] OR "puberty"[Title/Abstract] OR "young*"[Title/Abstract] OR "Child" [Mesh] OR "child*" [Title/Abstract] OR "Pediatrics" [Mesh])
